# An adjustable magnetic levator prosthesis for customizable eyelid reanimation in severe blepharoptosis: Design and proof-of-concept

**DOI:** 10.1101/2023.02.22.23286215

**Authors:** Nish Mohith Kurukuti, Melanie Nadeau, Eleftherios I. Paschalis, Kevin Houston

## Abstract

**Purpose:** Blepharoptosis is a common oculoplastic condition defined as incomplete opening of the upper eyelid. Although surgical approaches have become the mainstay for correction, they often fail to improve blink function. The purpose of this study was to develop an adjustable magnetic levator prosthesis (aMLP) as a non-surgical treatment option for severe ptosis, that allows blink reanimation.

**Methods:** Magnetic force required to perform blink reanimation was characterized by evaluation of eye opening and closing using inter-palpebral fissure (IPF) outcomes with various combinations of eyelid array magnet and box magnets. The force that provided the optimal eye opening without limiting eye closing was selected for the development of the aMLP. The aMLP included an eyelid array magnet and an adjustable rotating spectacle magnet that allowed change in the magnetic direction, and hence change in the net magnetic interactive force between the spectacle and lid magnets. The clinical feasibility of the aMLP in improving eye opening without limiting eye closing was evaluated in ptosis patients through a proof-of-concept study using IPF and comfort outcomes.

**Results:** Optimal eye opening and closing was achieved by a magnet-array combination providing 45gF in the tested ptosis population. Optimal size of spectacle magnet for aMLP was selected based on the simulations of rotating diametrically magnetized cylindrical magnets to achieve force ranges required for blink reanimation. The aMLP was able to modulate eye opening and closing with change in rotation (fitting) of the spectacle magnet in two ptotic patients. The best fitting of aMLP improved IPF opening without limiting eye closing (spontaneous or volitional blinking) with good comfort reported.

**Conclusions:** Preliminary results suggest that the aMLP can correct ptosis without adversely affecting blink function. Further evaluation in a larger patient population is warranted.

**Translational Relevance:** A non-surgical, proof-of-concept, adjustable magnetic treatment option for blink reanimation in patients with severe ptosis is presented.

## INTRODUCTION

Blepharoptosis (Ptosis) is a common ophthalmologic condition defined either as unilateral or bilateral incomplete opening of the upper eyelid (drooping of eyelid) (Johnson 1964). It occurs due to abnormalities in the function or structure of the levator palpebrae superioris muscle responsible for lifting the eyelid. The etiology for ptosis can be divided into structural, restrictive, and paralytic. Paralysis of the levator muscle may result from disease of the 3^rd^ cranial nerve or dysfunction of the neuromuscular junction (Ahmad, Wright et al. 2011). Ptosis is prevalent in all age groups and requires treatment when it is severe and the visual axis becomes blocked. If untreated, it may lead to obstruction of vision in adults, or maldevelopment (i.e. amblyopia) in children (Patel, Carballo et al. 2017).

Current treatment of severe paralytic ptosis is suboptimal. Most patients are treated with the frontalis sling(Leatherbarrow 2010), which employs the frontalis muscle to compensate for the loss of levator muscle function. This treatment requires conscious effort by the patient to blink and does not allow normal or complete blinking (Nadeau, Houston et al. 2020). This can lead to substantial risk for chronic exposure of the ocular surface (over-correction), which can be avoided by a more conservative approach, which typically results to insufficient eye opening.

In most types of ptosis, although opening the eyelid is impaired, the neuromuscular complex for eye closure remains intact (Leatherbarrow 2010). In these cases, an artificial levator prosthesis may be used to restore normal eyelid motility. To this end, previous studies have shown that re-animation of the levator muscle may be achieved by using electrical stimulation (Scott, Miller et al. 2019), muscle transposition (Zigiotti, Nesi et al. 2004), or artificial muscle (Senders, Tollefson et al. 2010). However, these are invasive procedures, irreversible and often lead to sub-optimal clinical outcomes. Alternatively, non-invasive approaches, such as taping of the eyelids open or propping the eyelid open with a wire on glasses (ptosis crutch) (Lapid, Lapid-Gortzak et al. 2000) can help in the short-term, but may also inhibit natural blink and lead to corneal damage. Moreover, the ptosis crutch approach requires continuous adjustment to maintain the eyelid elevation,(Conway 1973) which is cumbersome and can lead to corneal abrasion if not carefully performed.

More recently, Houston et. al. demonstrated successful implementation of a proof of concept, non-invasive, blink reanimation approach, using externally fixed static neodymium magnets to compensate for the loss of levator function (Houston, Tomasi et al. 2014). The device, referred to as the magnetic levator prosthesis (MLP), consisted of a spectacle frame mounted magnet and an eyelid array magnet embedded in polydimethylsiloxane (PDMS) elastomer which was attached to the ptotic upper eyelid using Tegaderm film. Appropriate fitting for eye opening without adversely affecting closing was obtained by adjusting the spectacle frames’ nose pads, and hence the distance between the two magnets, by adding buffers (physical barriers) to limit the approach of the two magnets or using different size magnets on the frame. Although preliminary results were satisfactory, laborious adjustments were required to fine tune the function of the device and considerable changes were required to fit the device between various patients. Moreover, while the MLP allowed better eyelid closing during volitional blinking than the ptosis crutch, it failed to allow natural spontaneous blinking in most of the participants.

To address the limitation of the MLP, we aimed to characterize the range of forces that would provide eye opening while not limiting spontaneous eye closing in the ptotic population. Following the characterization of the optimal range of forces for blink reanimation, optimal magnet dimensions to provide the desired forces over a range of distances that are feasible relative to MLP clinical use was determined. A Multiphysics model was generated to translate the results to parameters of the adjustable force. To improve the functionality of finer titration of magnetic forces in the selected range of the MLP and allow spontaneous blinking, we developed a new design with adjustable force dial on the side of the frame magnet casing. This allowed manual change of the magnetic axis of the spectacle magnet, thereby shifting of the direction of the magnetic lines and the interaction with the eyelid array. This arrangement is feasible and eliminates the need for electromagnets or electronics to alter the interactive force between the two magnets. Here, we employ this new design to test the hypothesis that angular translation of the magnetic field of the spectacle magnet modulates the force on the eyelid magnet in a manner that allows both volitional and spontaneous blink in a clinically feasible and useful manner. A prototype adjustable MLP (aMLP) was produced with 3-D printing, based on the magnet parameters determined in the force experiments and modeling, and tested with the help of two participants with severe paralytic ptosis. If our hypothesis was supported, it would demonstrate proof of concept of our novel angular translation approach while documenting the methods of prototyping the first ever adjustable force version of the MLP.

## METHODS

We created a bipartite system similar to that described by Houston et al. 2014, 2018, consisting of eyelid array magnet attached to the outer surface of the upper eyelid of the participant with ptosis, and a larger magnet attached to the spectacle near the eyebrow of the participant (spectacle magnet) (Houston, Tomasi et al. 2014, Houston, Tomasi et al. 2018). Eyelid array magnet was fabricated with 3 rectangular cube magnets of length 3mm, width 2mm, and height 1mm (Figure 1A) embedded in polydimethylsiloxane (PDMS). There were two polarization orientations of the arrays tested (Figure 1A): Type-I eyelid magnets had polarization with north and south poles oriented through the width so that when worn by the participant poles ran in the same direction as the gravitational plane, and Type-II eyelid magnets had north and south poles oriented through the height so that when they were worn poles ran perpendicular to the gravitational plane. The encapsulated magnets, referred to as the “array”, were attached to IV 3000 Flexifit (Smith and Nephew, UK), similar to those described by Houston et al. (Houston, Tomasi et al. 2014, Houston, Tomasi et al. 2018), with the exception that the PDMS array was bonded to the outside of the adhesive, rather than draping the adhesive over the array during application.

**Figure 1.**
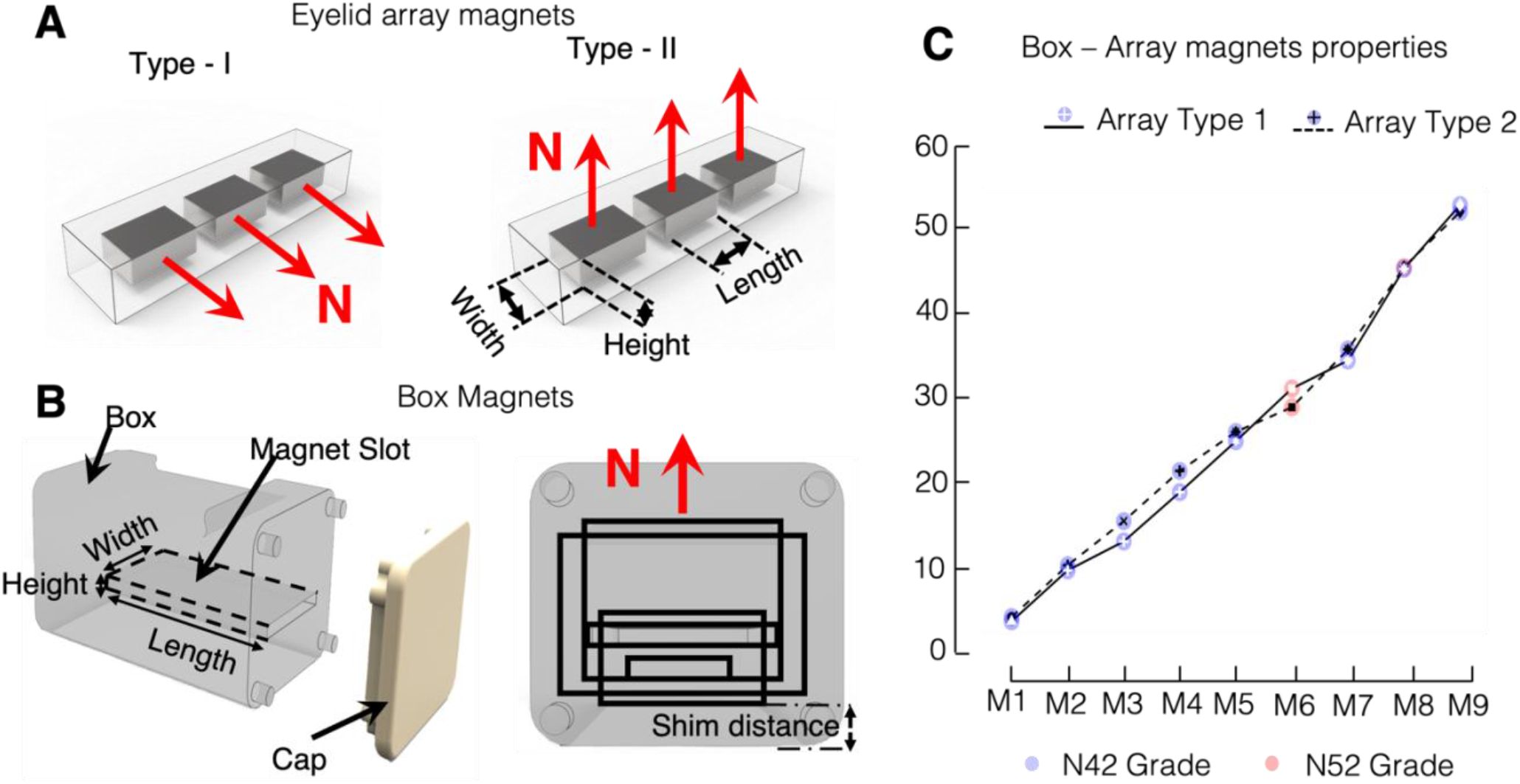
Experimental setup to study the effect of increasing magnetic force on blink dynamics (Box Experiment). **A**, Eyelid array magnets are small (L x W x H: 3 × 2 × 1mm) rectangular magnets embedded in PDMS that is bonded on IV 3000 skin adhesive film. As shown by the red arrows, Type-I arrays have the manufacturer-defined magnetization orientation along the height of the magnets while Type-II have magnetization orientation through the width (SM magnetics, Pelham AL, USA), dot marker on denoted pole. **B**, Boxes had a slot where rectangular static Neodymium grade 42 or 52 magnets (M1 to 9) were inserted to produce magnetic surface force ranging from 5 to 55gF in 5-10gF increments, verified with strain gauge. Shims were used to make minor adjustments in surface force as needed. All boxes (M1-9) looked identical and were arbitraily labeled to mask the operator and participant to size and force. The north pole of the box magnet was oriented upwards such that the north pole of the array was attracted to the bottom of the box. Black outlined rectangles represent the cross-sectional geometries of the various boxes. **C**, Surface pull force for each box. The y-axis represents empirical measurements of surface pull force in grams (gF) exerted on the type I and II arrays, measured with a strain gauge.

To characterize blink dynamics, we aimed to test a range of forces that would vary from having no effect on the eyelid (very weak) to very strong (where the eye could not blink). Non-custom rectangular NdFeB magnets of different sizes in grades N42 or N52 were obtained from SM Magnetics (Pelham, AL) or K&J Magnetic (Pipersville, PA), based on the manufacturer pull force specifications. Nine magnets (M1-M9), spanning a range of 5 to 55gf, with increments of 5gf between each magnet were selected (Figure 1B). To create more precise incremental forces without having to produce custom magnets, the magnets were inserted into 3-D printed boxes (Figure 1A) and “shims” of 1.5mm thickness were added until the desired surface full force was achieved, as measured with an array attached to a strain gauge. The 9 boxes were produced to appear identical, to mask the experimenter and participant to the strength of the magnets to attract the eyelid array magnets. The difference between the boxes was the size of the magnet housed within the box or the distance between the magnet and the testing surface of the box (Figure 1B). The boxes were prototyped with standard greyscale resins from Formlabs (Somerville, MA) and Stereolithography printing technology to obtain precise geometry of the 3D-printed boxes. Magnets were inserted into the boxes and sealed to prevent unmasking and boxes were numbered to facilitate randomization during testing. We heretofore refer to these as “box magnets”. Double masking (participant and experimenters) was also utilized for eyelid magnet polarization direction. The force range and increments were chosen based on unpublished preliminary tests which had the goal of providing a range beyond the physiological limits (beyond the force levels intended for clinical application).

### Box experiment

The change in magnet force using box magnets (M1 – M9) paired with eyelid array magnets (Type I and Type II) on blink dynamics was tested through a two-visit study design. In each visit, participant’s baseline eye opening, eye closing during volitional and spontaneous blinking, were measured without eyelid array magnets. Following baseline measurements, eyelid array magnets were placed on the ptotic eyelid of the participant after cleaning the eyelid with eyelid wipes. The experimenter held up different box magnets near the brow of the ptotic eye so that the surface of the box and arrays were in contact (see Fig1C). Next, participants were asked to blink normally (spontaneously) for ∼15 seconds. Afterwards they were asked to tightly close and open their eyes widely (volitional blink), and then to flutter their eyes (fast volitional blink). Presentation of box magnets to each participant was counterbalanced using the Latin Square method. Videos recording various blink types were recorded. Blink dynamics (eye opening, eye closing during volitional and spontaneous blinking) with inter-magnet distance (distance between eyelid array magnet and box magnet) were measured with different box magnets and eyelid array magnets combinations (Fig. 1A,B). Inter palpebral fissure measurements were taken from the videos of participants blinking with or without the magnets fitted. We applied mixed effect model analysis to the blink dynamics to evaluate the performance of box magnets for each eyelid array magnet type.

### Participant recruitment

All experiments were conducted in accordance with the declaration of Helsinki and approved by the institutional review board of Mass General Brigham (formerly Partners Healthcare). All individuals who participated in this study were recommended by referral of their physician. Participants were screened, enrolled, and met the inclusion criteria of having paralytic ptosis which obscured the visual axis without frontalis drive and the ability to provide informed consent or assent. Informed consent was obtained from all participants following detailed explanation of the nature and consequences of the study. Selection was not affected by age or gender. The authors affirm that human research participants provided written informed consent for the publication of the images.

### COMSOL modelling

To evaluate the concept of change in magnetic force through angular translation of the magnet and to select the magnet with operating range in the physiological range for clinical use, we developed a COMSOL model to perform electromagnetic simulations of the spectacle magnet and the eyelid array magnets. The model was validated experimentally for two conditions: change in inter-magnet distance between the eyelid array magnet and reference magnet and change in angular orientation of reference magnet with respect to the eyelid array magnet.

Validation of the model prediction of the change in force exerted on the eyelid array magnet by the reference magnet at various distances was performed using a strain gauge. The reference magnet, a 12.7mm × 12.7mm cylindrical diametrically polarized N52 magnet (SM magnetics, Pelham AL), was inserted in a rotatable housing similar to that in Figure 5A and mounted on a stand. A strain gauge (Grass Inc.) was mounted on a stand across from the reference magnet. A thin string attached the strain gauge to a 3-magnet eyelid array (both types were tested). The eyelid array magnet was placed into the field of the reference magnet such that their polarizations were opposite to each other, meaning the North pole of the eyelid array magnets faced the south pole of the reference magnet (opposing poles), but the two were not allowed to touch. The array was suspended in the field so that the string was taut. Next, the stand with the strain gauge was adjusted until the inter-magnet separation was 1mm, as measured with Vernier calipers. Using the same set-up, the stand with the strain gauge was adjusted to vary the inter-magnet separation. Force was measured at a separation of 1mm, 2.7mm, 5.7mm and 10mm. Similarly, the same environment with the eyelid array magnet and reference magnet was simulated in the COMSOL model and the inter-magnet distance was varied from 1 – 10mm, while the force exerted on the surface of the eyelid array magnet was measured. The empirical and simulated measurements were compared for correlation using Pearson’s correlation.

Likewise, validation was also performed on the predicted change in orientation of the reference magnet with respect to the eyelid array magnet. The same reference magnet was simulated along with the eyelid array magnet with inter-magnet separation of 10mm and the reference magnet orientation of 0° (opposing poles). The reference magnet was rotated in 45 degrees increments from 0 to 180 while the force was measured from the surface of the eyelid array magnet. Empirical force measurements were also recorded with the strain gauge setup replicating the simulated conditions. The empirical and simulated measurements were compared for correlation using Pearson’s correlation.

Upon successful completion of the validation, we evaluated candidate spectacle magnets dimensions for use with the adjustable MLP (aMLP). We simulated three diametrically magnetized cylindrical magnets of diameters 12.7 mm (12.7D), 9.53 mm (9.53D), 6.35 mm (6.35D) with the same length of 12.7 mm. All magnets were simulated with the type-II eyelid array manget. The simulations involved calculating the force on the surface of the eyelid array magnet with varying the inter-magnet distance from 3-22 mm and with orientation of the spectacle magnet from 0 - 180°.

### Adjustable Magnetic Levator Prosthesis

Three prototypes were produced, tested, and refined in this testing process (Supplementary figure 2). The final design was produced and used in the proof-of-concept human studies. Solidworks CAD (Waltham, MA) was used to design a spectacle clip-on and magnet casing with compliant dial system that was 3-D printed and attached to a frame (most visible in Figure 5). The magnet case was a separate piece that slid onto a rail on the clip-on. The clip-on, most easily visible in Figure 5, had a base, which was designed to follow the contour of the frame for stability when attached onto the frame using clips and brackets. It consisted of two rails on either side of the frame nose bridge, with triangular ridges, onto which the magnet holder was mounted. The magnet holder was designed to rotate the cylindrical magnet by 360 degrees with 30 degrees steps using a simple rocker-pinion mechanism. It was mounted onto the clip-on base at the rails using the clamp. The clamp had a triangular ridge that meshed with the triangular ridges on the rails to restrict the motion of the magnet holder. The magnet holder had a cavity to house the circular spectacle magnet. The spectacle magnet was glued onto the holder dial that had a gear with its teeth 30 degrees apart from each other. The teeth of the gear meshed with the monolithic stopper in the magnet holder, which facilitated the rotation of the dial by 30 degrees steps. It restricted the rotation when force was not imparted onto the dial. We analyzed the design for stresses and flexibility that the rotation mechanism could withstand with various 3D printable materials using Solidworks (Waltham, MA). Following the analysis, we selected Polyamide 11 (Nylon 11) as the material to 3D print the clip-on, magnet holder and holder dial using Selective Laser Sintering (SLS) printing technology (CR 1989).

The effect of rotating the spectacle frame magnet in combination with eyelid array magnets (Type I and Type II) on blink dynamics was measured to show a proof of concept of an aMLP. Two Participants with severe ptosis (S6 and S10) were fitted the eyelid array magnet (Type-II) and aMLP frame prototype. Video recordings of the participants natural blinking were used to measure eye opening, and the amount of eye closing during volitional and spontaneous blinking at baseline and at various aMLP spectacle magnet rotations (0, 30, 60, 90, 180 deg) through interpalpebral fissure measurements. Fifteen seconds of recording from each setting was processed and analyzed (resulting in 250 image stacks). Comfort ratings were also recorded on a Likert-type scale of 1 – 10, with 10 being the most comfortable.

#### Data Processing and Statistical Analysis

Participants had interpalpebral fissure measurements during both experiments: box and AMLP experiments. In both experiments, participants were comfortably seated in a chair and videos of the participant’s blinking while looking straight ahead were captured using an iPhone X camera placed 50 cm in front of the eyes. Interpalpebral fissure (IPF) height between the bottom eyelid margin to the top eyelid margin through primary corneal reflection, was measured using ImageJ from the 30Hz iPhone video recordings with the eye open (eye opening) and at maximum closure during blinks (completeness of blink). IPF measurements were calibrated using the adult population norm 11.67mm white-to-white scleral distance, also referred to as horizontal visible iris diameter (HVID). HVID only varies approximately 0.26mm with sex and race (Matsuda, Woldorff et al. 1992). Eye opening was separated from blinks using an algorithm that detected eye blink when the amount of visible sclera was below a preset whiteness threshold while the eye was open and determined blink rate. An observer validated the output by watching the videos and verifying that the number of visible blinks matched the output of the algorithm.

All statistical analyses were performed in R and STATA. The outcome measures data were transformed into normalized eye opening, normalized volitional eye closing, and normalized spontaneous eye closing by subtracting the respective measures from mean baseline. The transformed distributions were inspected for normality and sphericity using Shapiro-Wilk test and Mauchly’s test of sphericity, respectively. Three linear mixed effect models were fitted to the data, one for each of the outcome measures with force generated by each magnet box M1-M9 as fixed effects for each eyelid array magnet type, with random intercept of each participant. Post-hoc pairwise comparisons (t-tests) between Box Magnets were then performed, with Bonferroni method to correct for multiple comparisons. Comparison between array types of box magnets M8 and M9 were done using a linear mixed effect model. Linear mixed effect model with fixed effects of Magnets (M8 – M9), Arrays (Type I or Type II), the interaction between Magnets and Arrays, with random intercept and slope for each participant were fitted to the data. Post-hoc comparisons of the interaction between Magnets and Arrays were made using Bonferroni corrected t-tests.

To compare various magnet forces (box magnets and eyelid array combinations) on blink dynamics (eye opening and closing) for each participant we computed optimality scores, which represents how well a combination of box magnet and eyelid array can open the upper eyelid while not limiting eye closing in both volitional and spontaneous blinking. Optimality score is computed for each box magnet by taking the difference between the amount of eye opening and volitional or spontaneous eye closing obtained through the box magnet and is normalized by the maximum optimality score achieved for each participant (eq. 1). Optimality scores help to compare the impact of the magnetic force between the box magnet force and the different types of eyelid arrays on blink dynamics for each participant. Optimality scores were computed for each box magnet and eyelid array magnet combinations in which eye opening, volitional and spontaneous eye closing values were measured. Combinations that did not have at least one of the blink dynamic measurements were excluded from optimality computation.

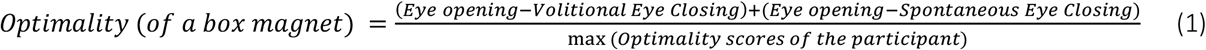

## RESULTS

### Participants

Fourteen participants met the inclusion criteria of paralytic ptosis which obscured the visual axis without frontalis drive and were enrolled after providing informed consent for a protocol approved by the institutional review board of Partners Healthcare. The study was conducted in accordance with the tenets of the Declaration of Helsinki. Three participants were excluded from analyses because they exited the study before data could be collected (2 due to early recovery and 1 due to scheduling conflicts & referral for a rigid scleral contact lens). Of the remaining eleven participants, median age was 55 years (interquartile range of 22) and 54% were female. Three had ptosis in the right eye, 5 in the left eye, and 3 had bilateral ptosis (Table 1).

**Table 1.** I Characteristics of the participants investigated.

| Participant | Age range (y) | Affected Eye | Severity | Sex | Etiology |
| --- | --- | --- | --- | --- | --- |
| S1 | 15 - 20 | Left | 6.5 | F | Cranial Nerve III injury, Brain abscess |
| S2 | 75 - 80 | Right | Total | F | Stroke, Nuclear Cranial Nerve III injury |
| S3 | 40 - 45 | Left | 6.4 | F | Cranial Nerve III injury, Tumor |
| S4 | 60 - 65 | Both | 5.1 | M | OculoPharyngeal Muscular Dystrophy |
| S5 | 55 - 60 | Both | 4.5 | M | OculoPharyngeal Muscular Dystrophy |
| S6 | 40 - 45 | Left | 5 | M | Congenital |
| S7 | 70 - 75 | Left | 0.6 | F | Sphenoid wing Meningioma |
| S8 | 50 - 55 | Right | Total pre-surgery | M | Chronic Progressive External Ophthalmoplegia |
| S9 | 80 - 85 | Left | Total | F | Cranial Nerve III injury, presumed Cerebro-vascular accident |
| S10 | 45 - 50 | Both | Partial (unspecified) | F | Chronic Progressive External Ophthalmoplegia |
| S11 | 65 - 70 | Right | Total | M | Stroke, Cranial Nerve III injury |

#### Blink Dynamics and Optimality scores from Box magnet experimentation

Linear mixed effect model analysis for normalized eye-opening (interpalpebral fissure, IPF) found significant main effect of box magnet force with both type-I (p < 0.001) and type-II (p < 0.001) arrays; therefore, as expected, more force led to more eye opening. Bonferroni corrected post-hoc t-tests revealed that M9 (55gF {grams of force}) provided significantly more opening than M1 (∼5gF), M2 (∼10gF) and M3 (∼15gF) (p < 0.001, t-test, Fig. 2A), but not M4-8 (∼20-45gF). This suggests that the opening effect may saturate at M4 (18gF) for the type I array. Post-hoc tests for type-II eyelid array magnets revealed a difference in IPF at M8 (∼45gF), being significantly greater than M1-7 (∼5-35gF), all P < 0.001, but not different from M9 (55gF). This suggests the opening effect may saturate at M8 (45gF) for the type II array. At 45gF (M8), the type-II eyelid array magnet provided significantly better eye opening than type I (p = 0.011, t-test, Fig. 2B), but at 55gF (M9), they were equivalent (p = 0.292, t-test, Fig. 1E). Therefore, results suggest that type II provides maximum opening with lower force.

**Figure 2.**
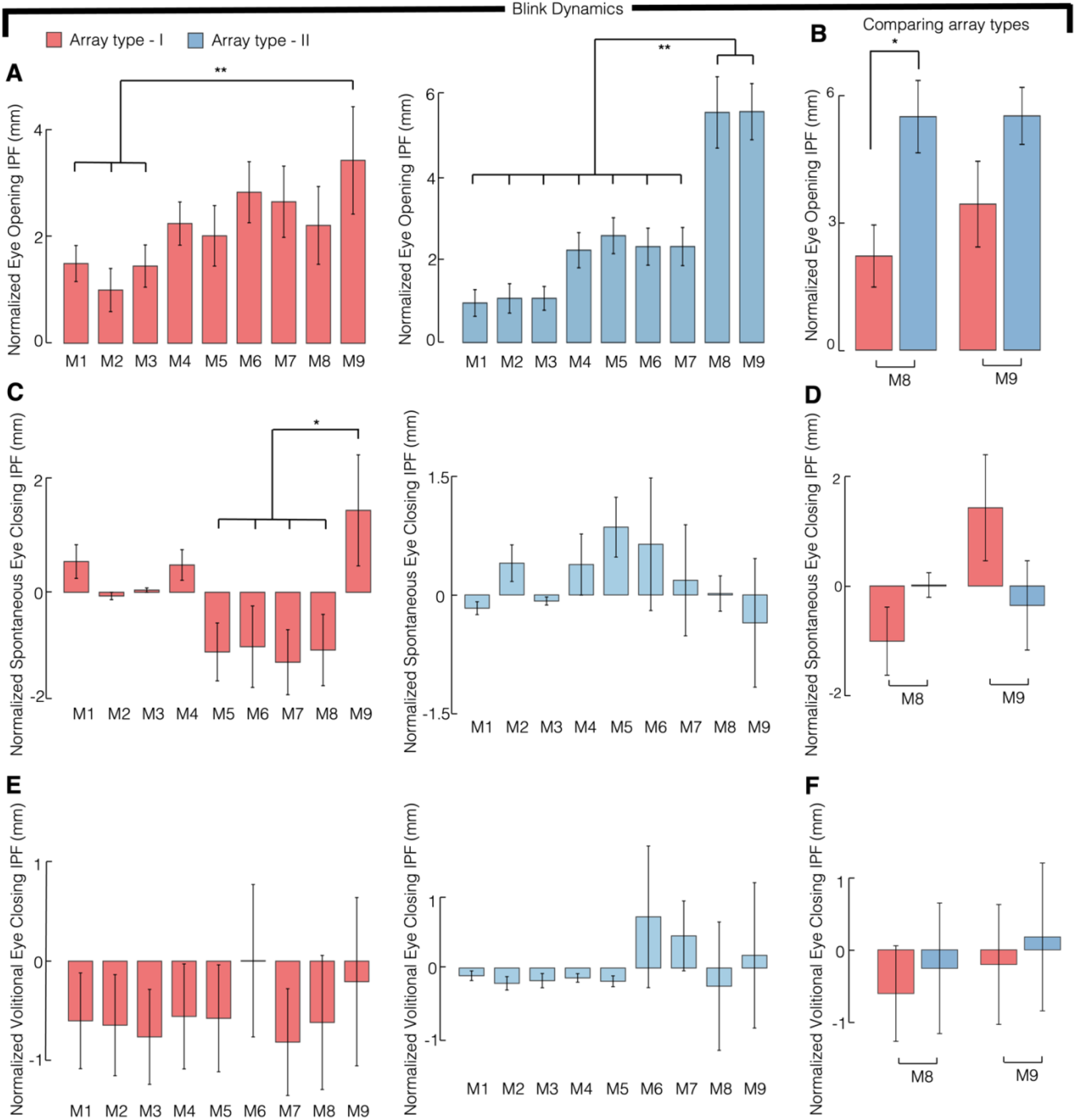
Results from characterizing eyelid dynamics with various box magnets and eyelid array type combinations. Bar plots of mean normalized values with standard error bars, where positive values represent increase in the interpalpebral fissure (IPF) height relative to baseline (without magnets) and vice versa. **A**, Magnets of any strength increased IPF, all bars >0), and higher force box magnets provided more eye opening. For type I (red bars), the 55gF magnet (M9) provided significantly more opening than lower force box magnets M1-3 (∼5-15gF). For type II (blue bars), M8 (∼45gF) and 9 (∼55gF) provided significantly more opening than lower force box magnets (M1-7, 5-35gF). **B**, Type-II array achieved significantly more eye opening than Type-I using M8 (∼45gF), without significant difference using M9 (∼55gF). **C**, Spontaneous blink was significantly impeded using M9 (55gF) for type I (red bars). For type II (blue bars), there was no significant relationship to force, suggesting the threshold was above 55gF. **D**, Spontaneous closing was not significantly different between type I and II arrays, for box magnets M8 (∼45gF) and 9 (∼55gF). **E**, None of the differences in volitional closing reached statistical significance; therefore, increasing the force as high as ∼55gF (M9) did not impede volitional blink for either array type.

Linear mixed effect model analysis for the normalized spontaneous closing IPF found significant main effect of box magnet force with type-I arrays (p = 0.017), but not type-II (p = 0.054). The lack of significance for type II suggests the spontaneous blink is not impeded at the range of forces tested (up to 55gF). For type I, Bonferroni corrected post-hoc t-tests revealed significant spontaneous blink impedance with M9 (∼55gF) (p < 0.05, t-test, Fig. 2C), with significantly worse blink IPF with M9 (∼55gF) than M8 to M5 (∼45-25gF). Pairwise comparisons of spontaneous closure IPF at M8 (∼45gF) and M9 (∼55gF) did not find significance for type I or type II arrays (Fig. 2F). Normalized volitional closing did not vary with increase in magnetic force. Analysis of the normalized volitional closing found no significant main effect for box magnets in type-I (p = 0.072) or type-II (p = 0.132) eyelid array magnet. Trends from both eyelid array magnet types show that with increased magnet forces volitional eye closing gets worse (Fig 2E). Comparisons between type-I and type-II eyelid array magnets of M8 (∼45gF) (p = 1.00) and M9 (∼55gF) (p = 1.00) also show that they were not significantly different from each other (Fig 2F). These results suggest that all combinations of box magnets and eyelid array types allowed a similar amount of volitional closing, which was very similar to the baseline volitional closing.

Data were normalized for each participant such that the best box magnet–eyelid array magnet type combination = 1. Optimality scores normalized for each participant showed different box magnets and eyelid array magnet type combinations being the best choice for various participants. Averaging the optimality scores across participants showed the combination of M8 with type-II eyelid array magnets (45gF) was the best overall option across participants (Fig 3). This suggests that M8 (45gF) with type-II array provides the overall best eye opening while not limiting eye closing during the volitional or spontaneous blink. However, it also shows that having customizability of magnetic force can be beneficial, because the best combination varied widely across participants (Fig. 3).

**Figure 3.**
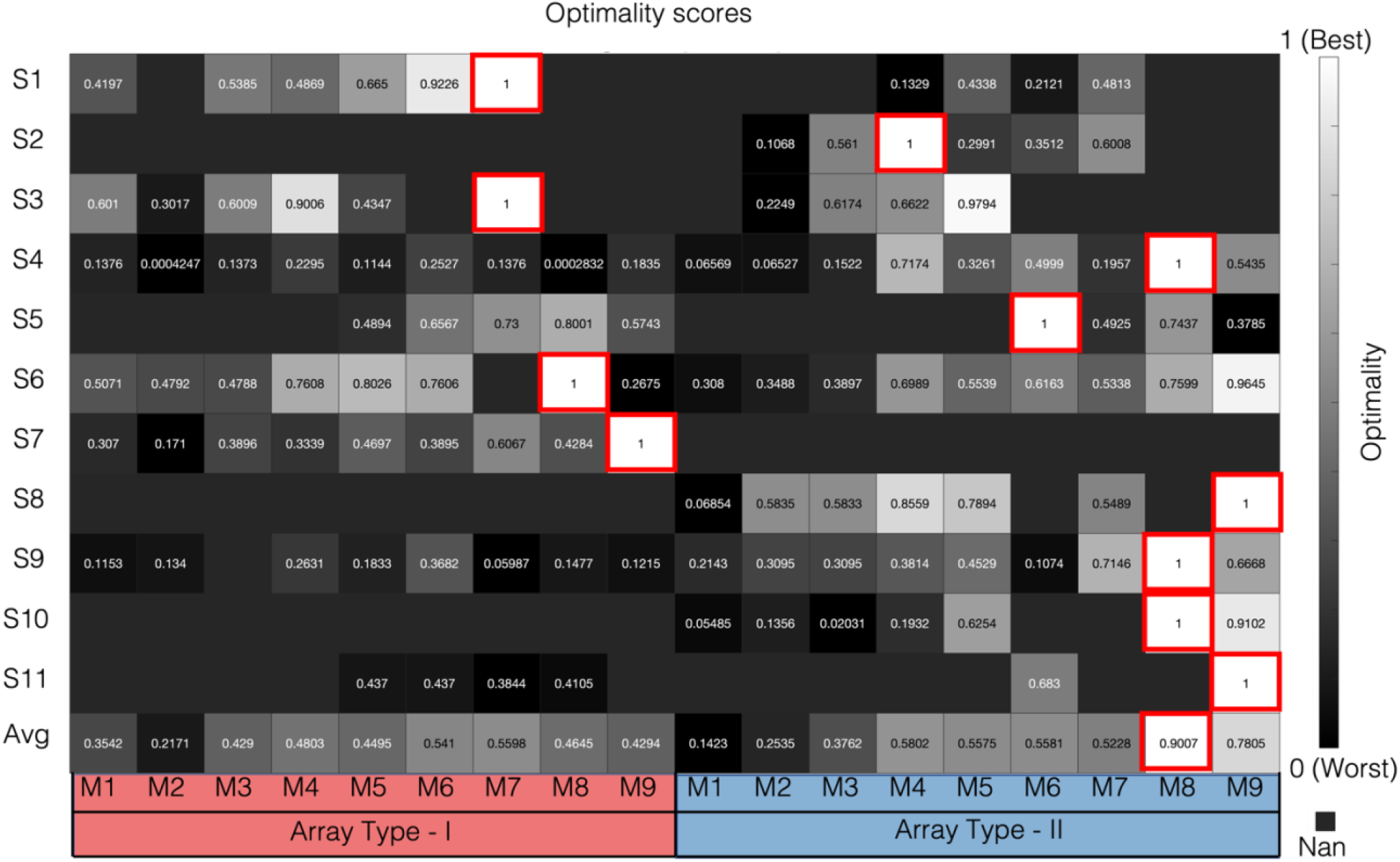
Optimality scores for each box magnet-eyelid array magnet combinations for all participants. Scores were normalized for each participant such that a score of 1 (red outline) was assigned to the box magnet eyelid array combination that provided the best eyelid dynamics for opening and closing (spontaneous and volitional). The blank areas in the chart represent missing data. Some participants could not complete all testing. The combination of box magnet M8 (45gF) with eyelid array magnet type-II had the highest frequency of optimality equal to 1 (n = 4). However, a wide variation was documented in the chart, suggesting the need for use of both array types and an adjustable spectacle magnet force from 18gF to 55gF (M4 to M9).

### COMSOL modelling

Based on the results of the box experiments, a range of 30gF (M6) to 55gF (M9), as measured at the box surface, could optimally fit >90% of the target population sample. This computed to 7-15gF at a clinically feasible inter-magnet separation of 8mm. We hypothesized that this range of force could be provided via angular translation of a static, diametrically polarized, cylindrical neodymium magnet of a size feasible to wear on a spectacle frame (e.g. < 15mm × 15mm). The adjustable force feature was realized using angular translation, i.e., the magnet on the spectacles was manually rotated using a dial with predefined angular steps of 30 degrees (Houston, Ilios et al. 2020). Using COMSOL, the design was simulated to verify empirical data using a strain gauge (see methods, magnetic force testing), which also allowed further optimization of the system using COMSOL.

The distance between the spectacle frame mounted magnet and type II eyelid array was systematically increased and the force acting on the eyelid array was measured. As can be seen in Figure 4A, empirical data confirm simulated results (Pearson’s r = 0.995; p < 0.001), and the upper end of the necessary range (15gF) was reached at a distance as high as 9mm using a 12.7 × 12.7mm diametrically polarized N52 cylinder (the reference magnet).

**Figure 4:**
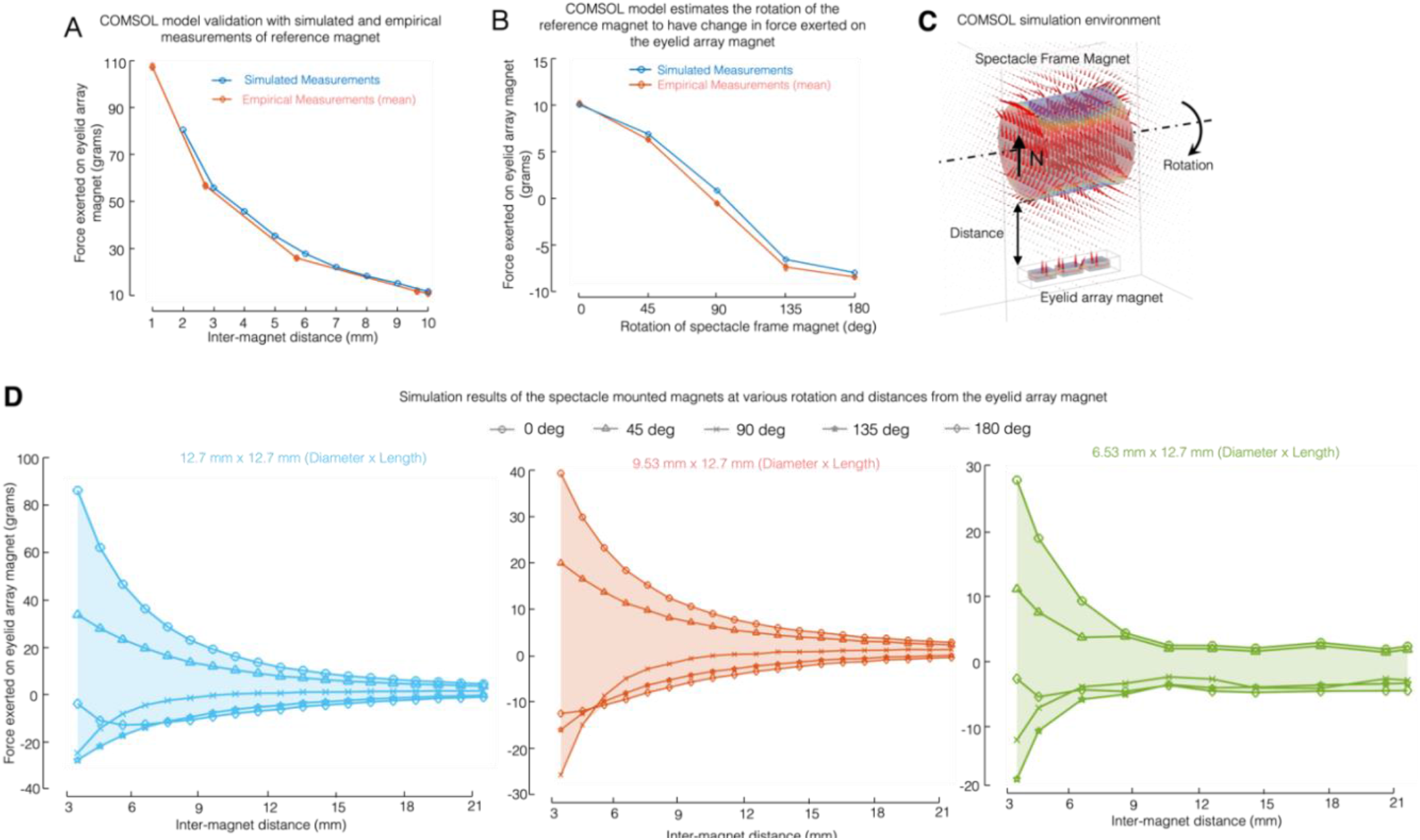
COMSOL model validation and simulated results for force modulation using angular translation, inter-magnet linear distance translation, and spectacle magnet diameter variation. **A**, Line plot showing minimal difference between empirical and COMSOL simulated force measurements. Measurements represent forces of a diametrically magnetized N52 reference magnet (12.7mm × 12.7mm cylindrical magnet) on a type II eyelid array. The eyelid array reference magnet were placed at various predefined distances [1mm – 10mm] (x-axis) and force acting on the top surface of the eyelid array magnets was measured 3 times each with a strain gauge (y-axis). **B**, Line plot showing minimal difference between COMSOL simulated and empirical force measurements of force variation (y-axis) with spectacle magnet angular translation (x-axis), validating the COMSOL model. Measurements represent the forces of a diametrically magnetized reference magnet with the type-II eyelid array magnet separated at a distance of 10mm. At 0° both magnets poles are aligned in the same direction (reference magnet south pole is aligned with eyelid array magnets north pole). The reference magnet was rotated by 45° steps from 0 to 180 degrees and force was measured 3 times each with a strain gauge. **C**, COMSOL model used for simulation of the environment of the spectacle frame magnet or reference magnet and the eyelid magnet array. The eyelid array was simulated as encapsulated in the PDMS substrate. The space between the eyelid array magnet and the spectacle frame magnet is simulated as air. **D**, Line plots reporting the results of simulations. Three magnet sizes were simulated while varying the angle of the spectacle magnet (y-axis) and inter-magnet distance(x-axis). Rotation angles were simulated at 45° steps. The shaded region represents the range of forces that can be exerted by the spectacle magnet on the type II eyelid array magnet due to a change in inter-magnet distance and angular rotation. Negative force values represent repulsion and positive force values represent attraction.

**Figure 5:**
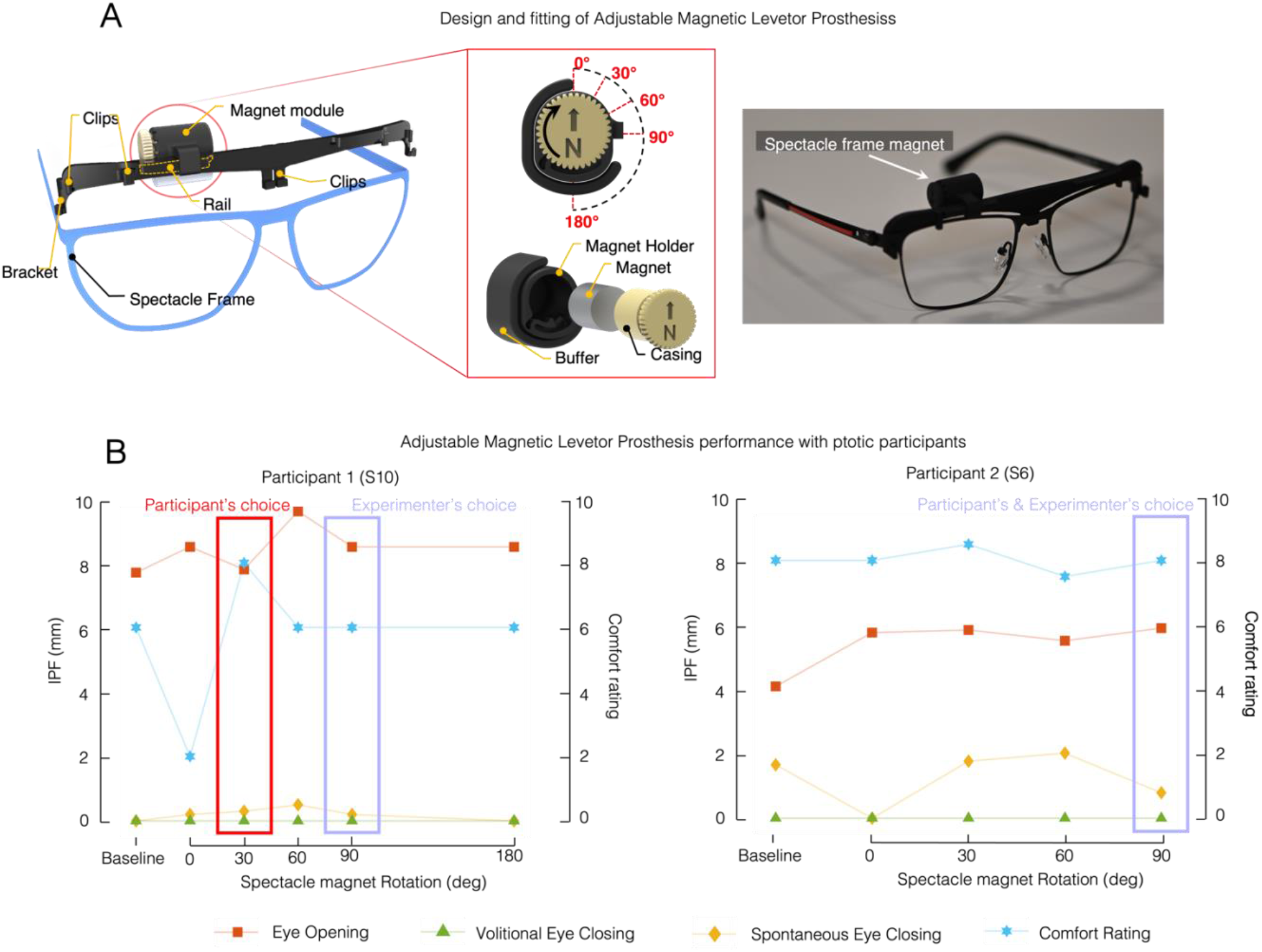
Conceptual and technical design of a prototype adjustable magnetic levator prosthesis and results of proof-of-concept testing for blink reanimation in 2 participants with severe ptosis. Adjustable magnetic levator prosthesis prototype generated for proof-of-concept testing in 2 participants with severe ptosis (S7 and S12). The prototype consisted of a 3D printed holder that attached to the spectacle frame (A). The spectacle frame magnet was mounted onto the clip and was manually adjustable laterally on a rail (outlined with dashed orange line), by the clinical study staff or participant, in order to position it above the eyelid array of the participant. The cylindrical 9.53×12.7mm N52 spectacle frame magnet was housed inside the magnet module (A, inset) and was designed to allow rotation (angular translation) in 30-degree steps to change the force exerted on the eyelid array magnet. (B) Line plots represent various blink dynamic parameters and comfort scores given by the participants for various orientation settings of the prototype aMLP. Participants and the examiner together chose the best setting.

As can be seen in Figure 4B, empirical and simulated measurements were essentially identical for various angles (Pearson’s r = 0.998; p < 0.001). The force exerted on the type II eyelid array magnet varied from -8 to +10gF. As shown in R2.1.1, the necessary upper range of 15gF should be met by reducing the magnet separation from 10 to 9mm, which could be easily accomplished in the clinic via nosepad adjustment. To confirm this and in the interest of making a device as small and lightweight as possible, we used the validated COMSOL model to vary size, distance, and angular orientation.

Three N52 diametrically magnetized cylindrical magnets with three different diameters of 12.7 mm (12.7D), 9.53 mm (9.53D), 6.35 mm (6.35D) but with the same length of 12.7 mm were simulated. Based on the results from the COMSOL simulations, the peak attraction force (positive values of force) provided by the magnets was proportional to the size of the magnet. The 12.7D magnet provided almost twice and three times the peak force compared to the 9.53D magnet and the 6.35D magnets. The range of force modulation possible with angular translation expanded dramatically when the magnets were closer together. For example, decreasing separation only from 8 to 7mm increased the range 4gF in the positive direction. The separation of the force lines can provide some guidance for the predicted fitting of the device. Namely, to get some potential effect of angular translation with the 9.5D mm magnet, results suggest that the inter-magnet distance should be no more than 12mm. Beyond this point there is very little (less than 1gF) separation in the force lines (see Figure 4D, plot lines for orientations 0 and 45 degrees). At a clinically feasible inter-magnet distance of 8mm, the 12.7D cylinder magnet provided 20gF with orientation of 0 degrees and -18gF for 90 degrees orientation, 9.53D cylinder magnet provided +12gF with orientation 0 degrees and -10gF with orientation 90 degrees, and 6.35D cylinder magnet provided 5gF with 0 degrees and -5gF with 90 degrees orientation (Figure 4D).

### Adjustable Magnetic Levator Prosthesis for Customizable Blink Reanimation

Based on the simulation results, 9.5 × 12.7 mm (diameter x length) was selected for initial prototyping (comparison of force exerted on the eyelid array magnet by various magnets are shown in Supplementary Figure 1). A prototype 3D printed frame clip-on was produced (See Methods, Adjustable Magnetic Levetor Prosthesis and Fig. 5A). This clip-on had a magnet module attachment, which housed the spectacle mounted magnet and facilitated change in orientation of the spectacle mounted magnet through a gear and monolithic stopper at steps of 30°. The range of force produced by the simulated magnet system ranged from 40gF at 3.6mm separation and 0 degrees angular orientation to -28gF at 0mm separation and 90 degrees of angular orientation. At a clinically feasible inter-magnetic distance of 8mm, angular translation of the spectacle magnet could vary the force on the array by 22gF (−10gF to +12gF). The prototype aMLP system, which had a magnet case plus buffer material of 3mm thick, had a max surface force of 40gF with type II array at orientation of 0 degrees. The box experiment suggested that the spontaneous blink would not begin to be impeded until forces reached 55gF (Fig. 2).

Next, the prototype was tested with two participants (S7 and S12, Table 1) with ptosis who also participated in the box experiment, in a protocol approved by the institutional review board. The participants wore the aMLP and were monitored with video recording for spontaneous and volitional blinks. We measured the interpalpebral fissure and comfort ratings on a Likert-type scale of 1 – 10, with 10 being the most comfortable. Both the participant and the examiner fitting the aMLP were asked as to provide efficacy ratings on a Likert-type scale of 1 - 10, with 10 being the most effective, for overall functionality provided by each orientation setting of the spectacle frame mounted magnet of the aMLP. Here we report the comfort and efficacy ratings for the best orientation setting chosen by the participants.

For the first participant (S10, top panel of Fig. 5B), wore the aMLP on the right eye and chose one of the lowest magnetic force settings (30°) as the best setting, while the clinical study staff chose the 90° orientation as the best setting. While both settings provided similar levels of eye closing (spontaneous and volitional), 90° setting provided better eye opening than the 30° setting. The participant rated the 30° setting higher (8/10) for comfort compared to the 90° setting (6/10). The inter-magnet separation with the best clinical staff fitting was 5mm when the eye was closed and 3mm when the eye was fully open, producing a COMSOL predicted force range of -6gf to - 20gf. However, despite the negative values suggesting repulsion, we noted that the array rolled over, reoriented the poles to produce attraction (See photos, Fig. 5B). The eye opening improved from 7.62mm at baseline to 8.65mm with the aMLP. The minimum spontaneous blink was 0.15mm with aMLP compared to the 0mm at baseline.

The second participant (S6) (Bottom panel of Fig. 5B) the clinical study staff, with the participant’s input, was successfully fitted using a type-II eyelid array magnet and chose the 90° orientation as setting for the spectacle frame mounted magnet which provided adequate opening with a natural appearing and comfortable blink (8/10 rating) (Fig. 2F). The inter-magnet separation with this best fit was 6mm when the eye was closed and 2 mm when the eye was fully open, producing a COMSOL predicted force range of -8gf to -28gF. Despite the negative values suggesting repulsion, we noted and it can be seen in the photos in Fig. 5B that the array actually rolled over, reorienting the poles to produce attraction. The eye opening improved from 4mm at baseline, to 5.9mm with the aMLP. The minimum IPF during spontaneous blink was nearly complete with the aMLP at 0.79mm, possibly slightly better than the 1.6mm measured at baseline. The participant gave this rotation position a comfort rating of 8/10 and efficacy rating of 7.5/10.

## DISCUSSION

We aimed to develop a Magnetic Levator Prosthesis with adjustable force to address the need for customization in the magnetic correction of severe paralytic blepharoptosis. The study results support the hypothesis that our novel approach of angular translation is clinically feasible and produces an adequate range of magnetic forces for real time titration of the force that controls the lid’s position. Clinical empirical data were validated mathematically using COMSOL multi-physics analysis, which allowed further optimization of the parameters of the device in a lab setting. Proof-of-concept was subsequently confirmed by fabricating a prototype device that was used in 2 participants with ptosis and was able to change the interpalpebral fissure height and the amount of closing during spontaneous blinking by changing the spectacle magnet angular position (Fig 5). To our knowledge, this is the first demonstration of successful blink reanimation using a proof-of-concept manually adjustable MLP device. This adjustable force feature should help address issues of incomplete spontaneous blinking, as identified in our prior work [12] and provide improved fitting and ease of force titration. This addresses a major limitation of using fixed magnets for blink reanimation (Houston, Tomasi et al. 2018) which are known to partially impede spontaneous lid closing.

The masked box experiment with the box magnet is the first to systematically study the effect of magnet force on eyelid dynamics in severe ptosis during eyelid re-amination. Our results demonstrate saturation of eyelid opening around 55gF for type I arrays and 45gF for type II arrays (Fig. 2A) without impeding volitional blink. Spontaneous blink was impeded at 45gF for type I arrays and >55gF for type II. This apparent difference between arrays may be related to higher force at the magnet interface due to greater thickness of the type I array magnets in the polarized direction (2mm in type I vs. 1mm in type II). The force is also distributed over a wider area (6mm^2^ in type II vs. 3mm^2^ in type I). Despite this apparent difference, analysis did not find significance, i.e. differences between eye closing during spontaneous blinking was not different for type-I compared to the type-II eyelid array magnet, in this small sample. Analysis of the optimal settings showed that both type I and type II arrays should be available for fitting in future studies.

While the aMLP provided good blink dynamics in these two cases, there are likely ways to make further improvements. We observed that in these 2 cases with unilateral ptosis, where the spectacle frame magnet was fitted only on one eye, the lightweight frame that we selected would sag onto one side due to the weight of the spectacle frame magnet. Substantial force changes occur with frame movements as small as 1mm, therefore a more stable frame is paramount. This can be accomplished using custom designed 3D printed spectacle frames, specifically designed for the individual user, as reported previously in a recent phase I study (Houston and Paschalis 2022).

An interesting observation in this proof-of-concept study was the dynamic rotational behavior of the eyelid array when the spectacle frame magnet was set at 90° and 180° rotation. From COMSOL modelling, we expected that these orientations of the spectacle frame magnet would repel the eyelid array magnet as both magnets were fixed in the simulated COMSOL environment. However, in actual testing with the 2 cases reported, the eyelid array magnet rolled on the participant’s coronal axis, folding into the eyelid skin, effectively flipping the orientation of the magnetic poles so they aligned for attraction (0° rotation). The eyelid skin thickness likely affected the amount of rolling. Further investigation is necessary to determine inter-subject variability in this rotational response and how this characteristic may be harnessed to improve performance and cosmesis. For example, rolling of the array had the effect of hiding it within the lid skin, and this is cosmetically preferable. In addition, rolling of the skin around the array appears to improve lid response to magnetic pull, and acts similarly to blepharoplasty, where the lid is shortened to correct ptosis. One possible method to control the rolling could be to increase or decrease the amount of PDMS flange on the surface of the IV 3000. To this end, a COMSOL model should be developed to account for changes in orientation with resistance factor, such as degree of skin laxity and thickness. This will accelerate prototyping, reduce cost and remove the burden of testing with participants.

Our study has some limitations. In the box experiment, boxes were held in place by the experimenter. While this was a rapid, and therefore, clinically feasible way to test the range of magnet strengths, it may have allowed the experimenter to realize the size of the magnet in the box or help the patient blink by unintentionally moving the box downward. Future studies should use an interchangeable holder on a sturdy spectacle frame to eliminate this. Precise positioning of the box using landmarks, such as the eyebrow, should be used so that placement is consistent between and within subjects. Characterization of spontaneous versus volitional blinks was a potential problem in the study and is currently our best explanation for the bimodal distribution seen in the box experiment data. We used verbal instruction (blink rapidly = spontaneous, and close tightly and open as volitional). In fact, the blinks classified as spontaneous may not represent true spontaneous blinks. If this is the case, our results may under-estimate the force where true spontaneous blink would be affected. Video recording for longer durations without specific instruction to blink may have allowed us to capture true spontaneous blinks, and if so, the results could be different. We only chose box magnets in the range of 5 – 55gF due to the lack of availability of off-the-shelf rectangular magnets. To know the exact upper limit for the optimal range of magnetic force, higher magnetic force testing would be required. With higher magnetic forces, we expect that eye opening would reach a ceiling while it would eventually completely prevent eye closing (volitional and spontaneous). This would be painful, however, and may not be ethical (or necessary). Despite these limitations, the results provide an important proof-of-concept on the relevance of angular translation of the spectacle magnet to provide the opportunity for better fitting and ease of customization of the aMLP for blink reanimation in participants with ptosis.

## Conclusion

The aMLP prototype developed addresses the need for customization in correction of severe paralytic blepharoptosis by providing eye opening while not limiting eye closing. The study results support the hypothesis that the novel approach of angular translation produces adequate range of forces for custom titration of force at clinically feasible magnet sizes and distances. The proof-of -concept was confirmed with a prototype device in vivo with 2 participants with ptosis, demonstrating changes in interpalpebral fissure when changing the spectacle magnet angular position. This adjustable force feature addresses the limitation of using a fixed magnet on the MLP, which partially impeded effective spontaneous lid closing. These results warrant further evaluation in a larger patient population to determine the efficacy of the aMLP in restoring eyelid opening with maintenance of the blink.

## Data Availability

All data produced in the present study are available upon reasonable request to the authors

## ACKNOWLEDGEMENTS

The authors would like to thank Renita Sebastin, OD for participating in data collection of the box experiment and aMLP experiment.

## REFERENCES

Ahmad, K., M. Wright and C. J. Lueck (2011). “Ptosis.” Practical Neurology 11(6): 332–340.

Conway, J. S. (1973). “Alleviation of myogenic ptosis by magnetic force.” Br J Ophthalmol 57(5): 315–319.

Cr, D. (1989). Method and apparatus for producing parts by selective sintering. United States, University of Texas System. 4,863,538.

Houston, K., E. P. Ilios and M. Tomasi (2020). Active magnetic prosthesis for eye-lid re-animation, Google Patents.

Houston, K. E. and E. I. Paschalis (2022). “Feasibility of Magnetic Levator Prosthesis Frame Customization Using Craniofacial Scans and 3-D Printing.” Transl Vis Sci Technol 11(10): 34.

Houston, K. E., M. Tomasi, C. Amaral, N. Finch, M. K. Yoon, H. Lee and E. I. Paschalis (2018). “The Magnetic Levator Prosthesis for Temporary Management of Severe Blepharoptosis: Initial Safety and Efficacy.” Transl Vis Sci Technol 7(1): 7.

Houston, K. E., M. Tomasi, M. Yoon and E. I. Paschalis (2014). “A Prototype External Magnetic Eyelid Device for Blepharoptosis.” Transl Vis Sci Technol 3(6): 9.

Johnson, C. C. (1964). “BLEPHAROPTOSIS.” International Ophthalmology Clinics 4(1): 125–156.

Lapid, O., R. Lapid-Gortzak, J. Barr and L. Rosenberg (2000). “Eyelid crutches for ptosis: a forgotten solution.” Plast Reconstr Surg 106(5): 1213–1214.

Leatherbarrow, B. (2010). Oculoplastic surgery, CRC Press.

Matsuda, L. M., C. L. Woldorff, R. T. Kame and J. K. Hayashida (1992). “Clinical comparison of corneal diameter and curvature in Asian eyes with those of Caucasian eyes.” Optom Vis Sci 69(1): 51–54.

Nadeau, M., K. Houston, E. Paschalis, R. Sebastin, N. M. Kurukuti and p. thiagarajan (2020). Comparing Surgical to Non-surgical Magnetic Correction of Severe Blepharoptosis: Preliminary Results.

Patel, K., S. Carballo and L. Thompson (2017). “Ptosis.” Disease-a-Month 63(3): 74–79.

Scott, A. B., J. M. Miller and T. C. Namgalies (2019). “Activating the levator to elevate the eyelid.” J aapos 23(4): 219.e211-219.e214.

Senders, C. W., T. T. Tollefson, S. Curtiss, A. Wong-Foy and H. Prahlad (2010). “Force requirements for artificial muscle to create an eyelid blink with eyelid sling.” Arch Facial Plast Surg 12(1): 30–36.

Zigiotti, G. L., F. Nesi, L. Scorolli, A. Meduri and S. Vismara (2004). “Transposition of the levator muscle suturing it to a frontal sliding flap, using a single superior lid crease incision, in blepharoptosis.” Investigative Ophthalmology & Visual Science 45(13): 264–264.

