## Supplementary figures for "An adjustable magnetic levator prosthesis for customizable eyelid reanimation in severe blepharoptosis: Design and proof-of-concept"

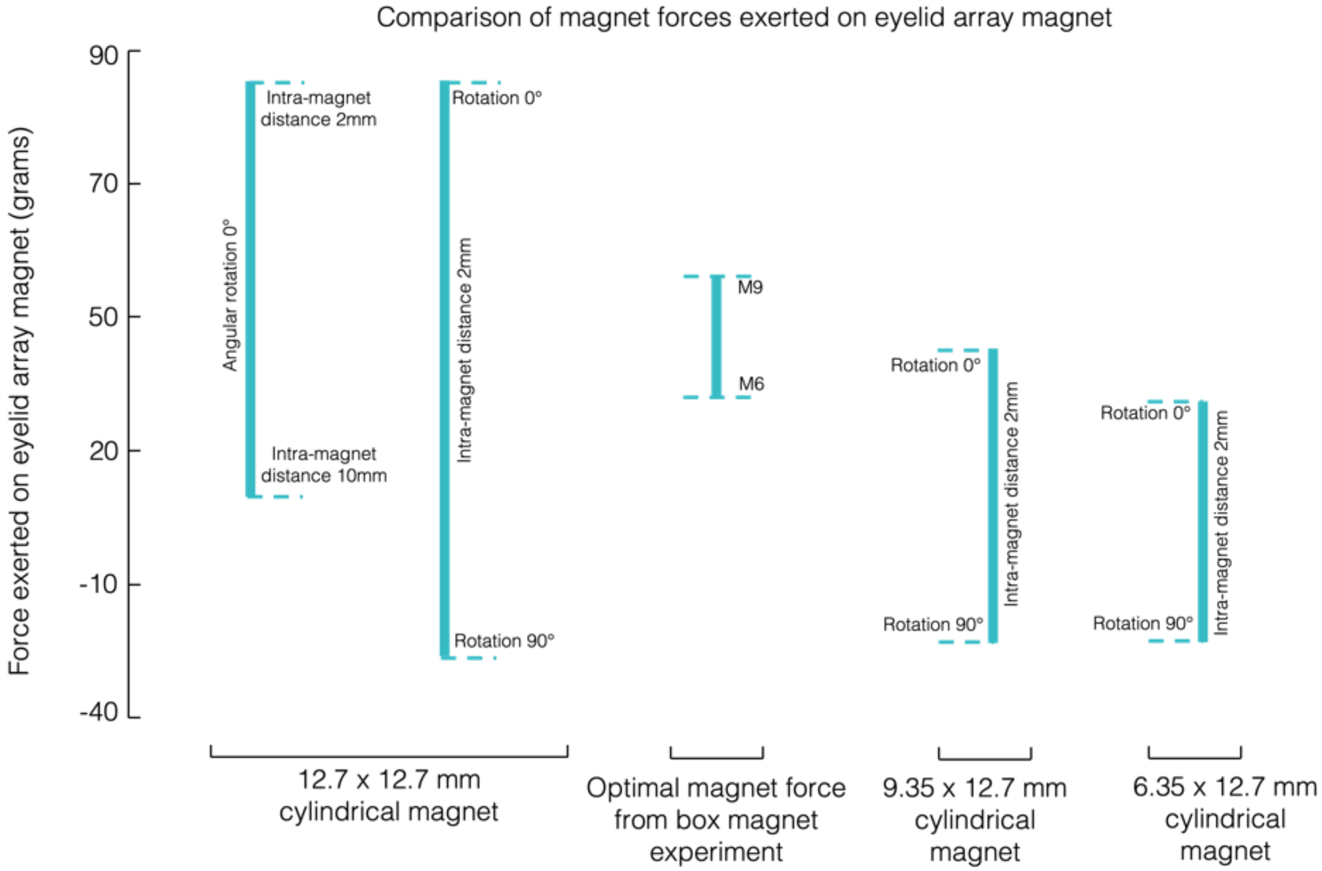


**Supplementary Figure 1 | Magnet force ranges for various magnets.** The reference magnet (12.7 x 12.7 mm) was empirically measured, varying the inter-magnet distance while keeping at the same orientation, and was also simulated using COMSOL by varying the orientation while keeping the inter-magnet distance constant. The other two magnets (9.53 mm x 12.7mm and 6.35mm x 12.7 mm) were rotated at various orientations while keeping the inter-magnet distance constant. These are plotted with respected to the peak force requirement from the box experiment.

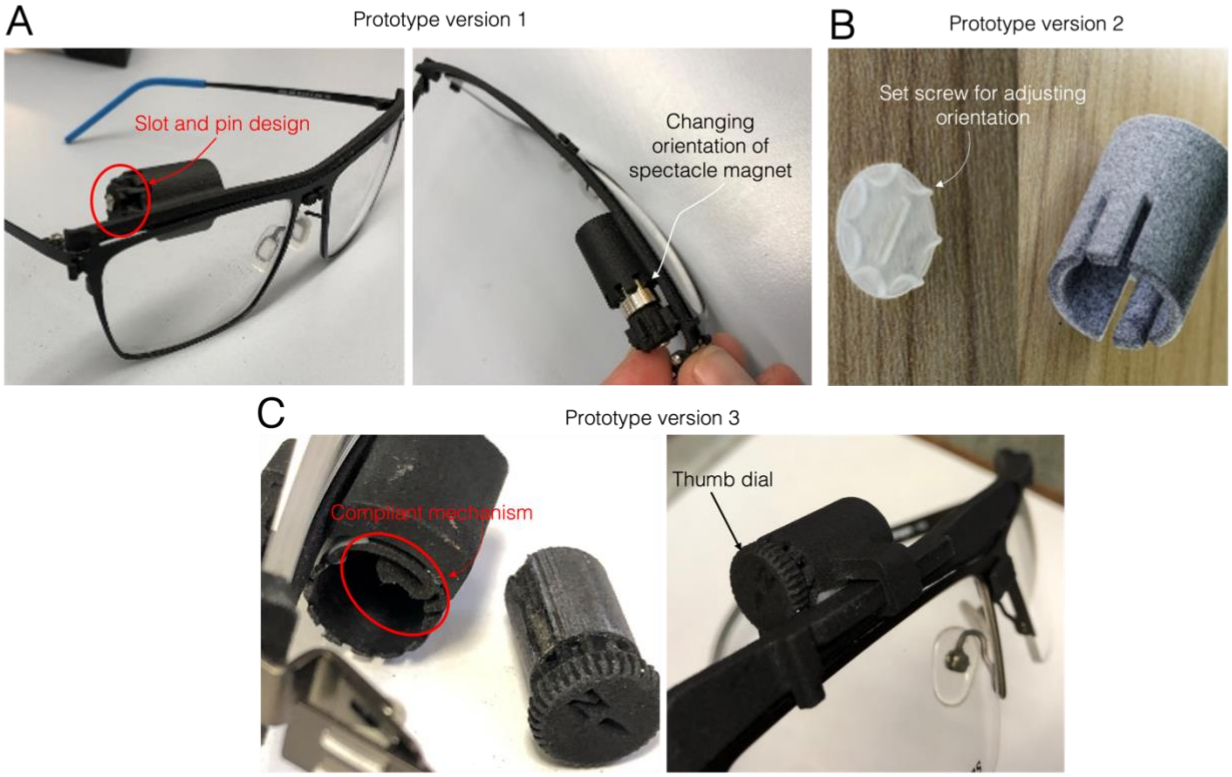


**Supplementary Figure 2 | Prototyping process for the aMLP rotatable spectacle mount**. A) First Prototype: A slot-and-pin design was employed. In order to rotate the magnet had to be pulled out, turned, and re-inserted. This exposed the magnet and was rejected by the clinical staff who felt it would be difficult for patients to self-adjust. In addition to the problems with the magnet clip, the frame was too flimsy, causing it to tilt from the weight and attractive force between the lid array and spectacle magnet, rendering poor efficacy. The magnet case was fused to the clip and could not be adjusted in the horizontal direction. This was a problem in fitting the participant as the spectacle magnet was not able to be centered over the eye. B) Second prototype: In this iteration, the magnet was drilled through its axis and a set screw attached it to the magnet case. The dial was glued to the surface of the cylindrical magnet. This approach was abandoned before testing with participants due to cost of custom drilling and negative effect of drilling on the magnet integrity. C) Third Prototype: This is the prototype that was ultimately able to demonstrate proof-of concept. The magnet was glued inside the compliant mechanism cap sleeve I, and then inserted into the case on the clip. The magnet could be rotated with thumb and index finger. A sliding rail allowed horizontal adjustment of magnet position. A sturdier frame was utilized and successfully maintained a more stable inter-magnet distance when compared to the Prototype 1 frame (A).
